# Navigating the Nexus of Food Insecurity, Anxiety, and Depression in the Face of Climate Change: A Longitudinal Study in Rural Kenya

**DOI:** 10.1101/2023.11.13.23298460

**Authors:** Michael Goodman, Lauren Raimer-Goodman, Heidi McPherson, Shreela Sharma, Ryan Ramphul, Dawit Woldu, Fridah Mukiri

## Abstract

**Objective:** To investigate the temporal relationships between food insecurity, anxiety, and depression among adult participants in a community-based empowerment program in Meru County, Kenya.

**Methods:** A cross-lagged panel analysis was conducted using data from 362 adult participants in a community-based empowerment program in Meru County, Kenya. Participants completed self-report measures of food insecurity, anxiety, and depression at two-time points, 11 weeks apart.

**Results:** Food insecurity (T1) predicted subsequent anxiety and depression (T2), controlling for within-variable, within-time, and control-variable correlations. Village-level food insecurity (T1) was correlated with significantly higher anxiety (T2). Additionally, anxiety (T1) predicted higher subsequent food insecurity (T2).

**Conclusion:** Food insecurity and anxiety have a complex bidirectional relationship. Interventions that address food security, mental health, and the psychosocial factors that promote adaptation to food-insecure environments are essential for promoting the well-being of individuals and communities in the face of climate change.

## INTRODUCTION

Climate change presents an increasingly urgent threat to human well-being through multiple pathways. One such pathway includes linkages between climate change and food production within smallholder farming societies such as within Kenya and across sub-Saharan Africa (Stutch, Alamo, & Schaldach, 2021). Through disruption to rainfall patterns, degrading soil quality, and high temperatures (Thornton, Jones, Ericksen, & Challinor, 2011; Kalisa et al., 2020; Yengoh & Ardo, 2020), crop yields have declined and are projected to continue declining within societies whose main food source is smallholder farms. Any effort to determine or project the toll of climate change to human health must be informed by mechanisms by which human health is impacted by food security – the state in which “all people at all times have both physical and economic access to sufficient food to meet their dietary needs for a productive and healthy life” (USAID, 1992). Food insecurity indicates an inability to achieve these conditions through socially acceptable means (Nanama & Frongillo, 2012).

In addition to the consequences of food insecurity to human development (Conceição, Levine, Lipton, & Warren-Rodríguez, 2016), there has been growing consideration of relationships between food insecurity and mental health (Weaver & Hadley, 2009). However, most current research assessing food insecurity and mental health depends on cross-sectional data from high-income countries; for example, the meta-analysis reported by Poumotabbed et al. (2020) relied on data from seven high-income countries, two upper-middle-income countries and one middle-income country. Further, the Pourmattabed et al. (2020) meta-analysis found only one longitudinal study comparing relationships between food insecurity and mental health (Pryor et al., 2016). Pourmattabed et al. (2020) report consistent associations between food insecurity and depression and stress, but inconsistent findings between food insecurity and anxiety. Cross-sectional survey data from 149 countries demonstrate associations consistent with dose-response relationships between food insecurity and worry – a central feature of anxiety (Jones, 2017), though this study was not included in the Pourtmattabed et al. (2020) review, presumably due to measurement differences. Tuthill et al. (2020) published findings showing that persistent food insecurity, but not HIV, is associated with subsequent depressive symptoms among perinatal women in Kenya. A 2020 synthesis of qualitative research across sub-Saharan Africa indicated depression is understood as resulting from multiple domains – including challenges in securing and providing food for family members and related interpersonal conflicts (Mayston et al., 2020).

The global prevalence of depression and anxiety have increased over the past two decades, and the COVID-19 pandemic added substantially to the global mental health burden (Santomauro et al., 2021). In 2019, an estimated 300 million people globally were living with some anxiety disorder, while 280 million people were living with a depressive disorder (GBD 2019 Mental Disorders Collaborators, 2022). In order to understand the effects of climate change on global mental health, longitudinal research is required to clarify the relationships between food insecurity and common mental disorders – including anxiety and depression.

While food insecurity may contribute to mental health disorders, little is known about the psychological characteristics that promote adaptation to food-insecure environments. Workforce mental health research has established that common mental disorders undermine worker productivity (Bubonya, Cobb-Clark, & Wooden, 2017); conversely, mental assets enable adaptive processes – for example, by promoting entrepreneurial activity (Tugade, Fredrickson, & Feldman Barrett, 2004; Su, Liu, Zhang & Liu, 2020).

### Study Aim

This study assesses cross-lagged panel relationships of food insecurity, depression and anxiety among adult participants (n=362) in a community-based empowerment program in Meru County, Kenya. We consider whether correlations between these variables are significant only at the individual-level, and whether correlational patterns are identified between village-aggregated variables.

## METHODS

### Geographical location

This study was conducted in Central and South Igembe subcounties of Meru County, Kenya, which is located just north of Mount Kenya. Food security during the study period are shown in Supplemental Figures 1a-b. At the start of the study period, acute food insecurity was considered stressed in the study locations, but had progressed to “crisis” levels according to the Famine Early Warning Systems Network, 2023).

### Study setting

This study used data from participants in a community-based empowerment program that has previously been shown to improve mental health, parenting behaviors, income, and reintegration outcomes for street-involved children and youth (Goodman et al., 2021a; Goodman et al., 2023a; Goodman et al., 2023b). The intervention combines group-based microfinance practices, leadership development, and a novel behavioral health curriculum to promote values-based growth (Goodman et al., 2023b). A previously developed index of lending group-affiliated interpersonal trust, norms of reciprocity, sense of belonging, and cohesion was shown to be cross-sectionally associated with lower food and water insecurity (Goodman, Elliott, Melby & Gitari, 2022). Survey-based questionnaire data were collected in February 2023 (T1) and May 2023 (T2). The mean (sd) amount of time between observations was 11.5 (4) weeks.

### Recruitment Practices

Program participants were recruited through a combination of active and passive recruitment. Active recruitment began at the village level, where families with children living on the streets or families engaged in HIV care at Ministry of Health clinics were identified (Goodman et al., 2023b). These index families then recruited 25-29 other families to join a microfinance group. Additional families joined the program through passive recruitment, mostly by word of mouth (Goodman et al., 2021b). Participants were therefore nested within groups, which were nested within villages. Ten villages were represented in the current study.

### Participant selection

Once groups were formed, program evaluation was conducted to track changes in key variables over the duration of active program participation. Participants in this study were randomly selected during routine group microfinance activities using a random number generator. All willing participants (97.8% of all participants) were invited to draw a folded piece of paper from an opaque bag, with “1” indicating selection and “0” indicating non-selection. Each established group within a village was invited to participate in the evaluation. There were 362 individual participants, across 11 villages, with paired T1 and T2 data in this study.

### Study design

This study used an interventional cohort design, with two data collection waves.

#### Measures

The survey questionnaire was developed in English, translated into the local language (Kimeru), and then back-translated into English to ensure accuracy and consistency. The survey was administered by a team of trained local experts from two Kenyan universities in the study county.

### Primary measures

This study examines the cross-lagged relationships between three primary variables: generalized anxiety, depression, and food insecurity. Responses to all primary variables are averages of item-level responses, which are continuous.

#### Generalized anxiety

Generalized anxiety was measured using the GAD-7, a 7-item measure that assesses how often respondents have experienced symptoms of generalized anxiety in the past two weeks. Generalized anxiety is a common mental health disorder marked by persistent and potentially debilitating worry, sustaining negative affect and contributing to dysfunctional psychobehavioral coping (Newman et al., 2013). The response scale is 4-point Likert-type, ranging from “not at all” to “nearly every day.” The GAD-7 has been validated in Kenya and other global populations (Goodman et al., 2022). The GAD-7 had excellent reliability in the present sample (α=0.87 at T1; and α=0.91 at T2).

#### Depression

Depression was measured using the 21-item Beck’s Depression Inventory-II (BDI-II; Beck, Steer & Brown, 1996). The BDI-II measures symptoms such as sadness, hopelessness, guilt, irritability, and difficulty concentrating. Each item is rated on a 4-point scale, and the total score is calculated by summing the ratings for all 21 items. A higher score on the BDI-II indicates more severe depressive symptoms. The BDI-II showed acceptable reliability in both time points (α=0.82 in T1; α=0.83 at T2). The BDI-II has been used in various cultures with strong predictive and discriminant validity (Bernard, Dabis & de Rekeneire, 2017).

#### Food insecurity

Food insecurity was measured using the Household Food Insecurity Access Scale (HFIAS; Coates, Swindale & Bilinsky, 2007), a nine-item scale that assesses the severity of food insecurity in households over the past four weeks. The HFIAS is one of the most widely used and well-validated measures of food insecurity in the world (Chakona & Shackleton, 2018). Each item on the HFIAS is rated on a 4-point scale, and the total score is calculated by summing the ratings for all nine items. A higher score on the HFIAS indicates more severe food insecurity. The HFIAS showed excellent internal reliability at T1 (α=0.94) and T2 (α=0.95).

#### Control measures

Age, estimated household monthly income, years of education, marital status, and gender (all from T1) were used as control measures. Age, income, and years of education were measured as continuous variables. Marital status (married/cohabitating vs. not married/not cohabiting) and gender (man or woman) were included as binary variables. All control variables have been found to correlate with depression, anxiety and food insecurity (Akhtar-Danesh & Landeen, 2007; Assari, 2017; Daig et al., 2009; Lee, Shin & Kim, 2020; Misselhorn & Hendriks, 2017).

#### Analytical approach

All scale variables were calculated by averaging item responses.

Descriptive data analyses include univariate descriptions of primary measures between T1 and T2. We used Wilcoxon signed-rank tests to evaluate equivalence of primary variables between T1 and T2. In bivariate analysis, Spearman rank correlation coefficients were calculated using Bonferroni-adjusted p-values to compare all primary and control measures.

At the individual-level, we conducted a cross-lagged panel analysis using Structural Equation Modelling (SEM) with robust standard errors, as some variables had distributions that were closer to Poisson than Gaussian. The preliminary model considered all pathways between T1 and T2 primary variables, and included all control variables in all pathways. Following a backward stepwise model-building approach, we recalculated the model until all variable pathways had significance tests p<0.20, and considered statistical significance at α<0.05. Figure 1 depicts this SEM-enabled cross-lagged panel model.

At the village-level, we analyzed correlations between village-aggregate primary values at T1 and T2 using Spearman rank correlation coefficient. Figure 2 depicts the resultant correlation matrix, Spearman rank correlation coefficient and respective p-value for the 11 included villages.

##### Ethical Considerations

All data were collected following ethical approval from the institutional review boards at the Kenya Methodist University and the University of Texas Medical Branch. The research was conducted under a research permit provided by the Kenyan National Commission for Science, Technology and Innovation (NACOSTI). All participants provided informed consent prior to engaging in the interviews. Following the preferred approach to compensation indicated by participants in interventional program, internal lending groups of each participant received $1 in exchange for their participation in the study.

## Results

Table 1, below, shows the univariate analyses of variables, and the probabilities that primary variables have equivalent values at T1 and T2. Food insecurity, anxiety, and depression all significantly declined between T1 and T2 (p <0.01 for each comparison). The average household monthly income was $35 at T1. The mean (sd) age was 41.7 (12.6) years. The mean (sd) years of formal completed school was 5.2 (3.2). The percentage of respondents who reported being married or living with a partner as though married was 76%, and 92% of respondents were women.

**TABLE 1:**
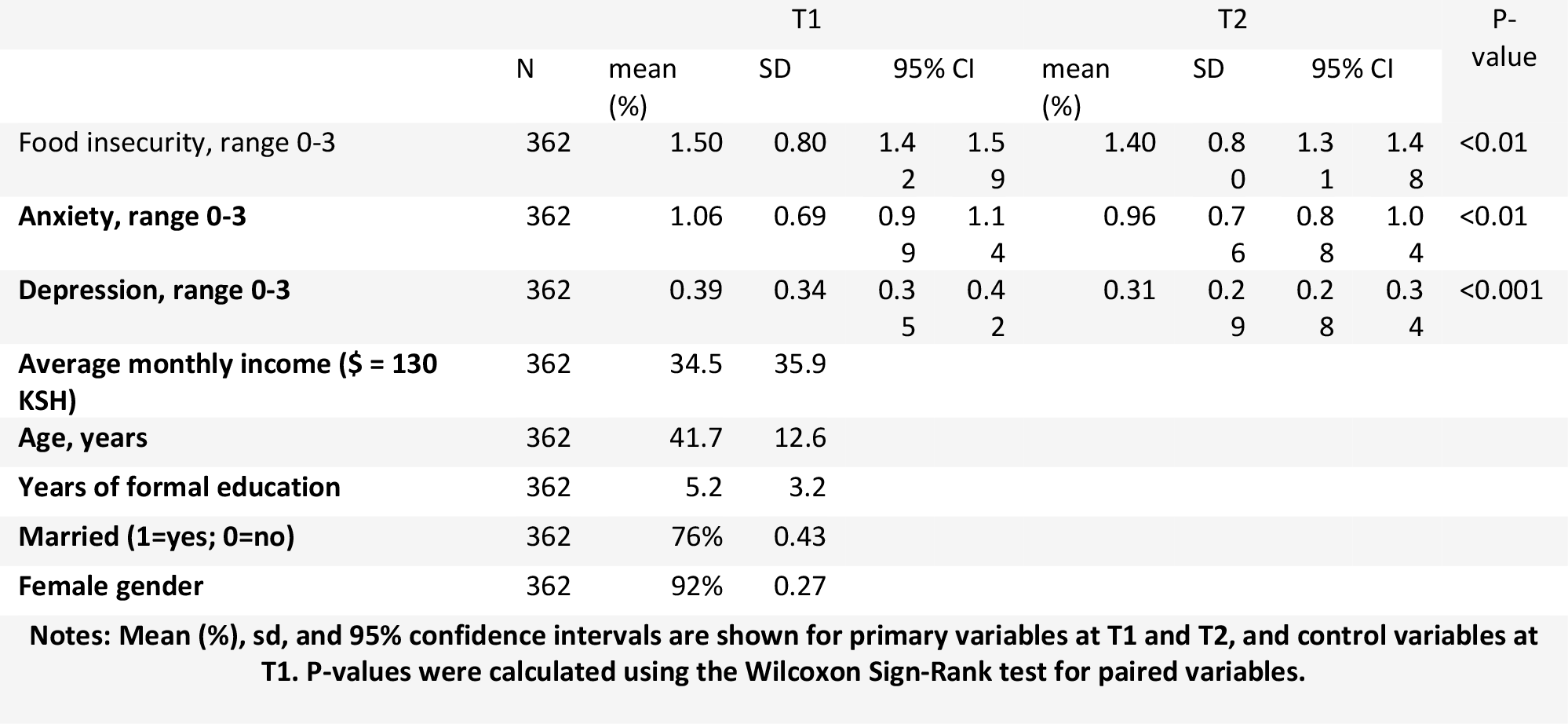
DESCRIPTION OF MODEL VARIABLES.

Table 2, below, shows the Spearman rank correlation coefficients with Bonferroni-adjusted p-values. Food insecurity (T1) was significantly correlated with all variables, except age. Food security (T2) was significantly correlated with anxiety (T1 and T2), depression (T2), and education (inversely, T1). Anxiety (T1) was significantly correlated with anxiety (T2), and depression (T1 and T2). Anxiety (T2) was significantly correlated with depression (T2). Depression (T1) was significantly correlated with depression (T2). Years of formal schooling were significantly correlated with higher monthly income and lower age (T1).

**TABLE 2:** SPEARMAN RANK CORRELATION COEFFICIENTS OF ALL MODEL VARIABLES.

|  | 1 | 2 | 3 | 4 | 5 | 6 | 7 | 8 |
| --- | --- | --- | --- | --- | --- | --- | --- | --- |
| 1. Food insecurity, T1 | 1 |  |  |  |  |  |  |  |
| 2. Food insecurity, T2 | 0.36*** | 1 |  |  |  |  |  |  |
| 3. Anxiety, T1 | 0.39*** | 0.22*** | 1 |  |  |  |  |  |
| 4. Anxiety, T2 | 0.24*** | 0.33*** | 0.29*** | 1 |  |  |  |  |
| 5. Depression, T1 | 0.26*** | 0.14 | 0.33*** | 0.15 | 1 |  |  |  |
| 6. Depression, T2 | 0.26*** | 0.27*** | 0.26*** | 0.49*** | 0.21** | 1 |  |  |
| 7. Monthly income, T1 | 0.36*** | -0.16 | -0.12 | 0.01 | -0.17 | -0.01 | 1 |  |
| 8. Age (yrs), T1 | 0.02 | 0.02 | -0.13 | -0.14 | 0.09 | 0.01 | -0.05 | 1 |
| 9. Years of formal schooling, T1 | -0.28*** | -0.36*** | -0.05 | -0.10 | -0.09 | -0.15 | 0.21*** | -0.35*** |

**Notes:** Spearman correlation coefficients shown of all primary (T1 and T2) and control (T1) variables. \* indicates p<0.05; \*\* p<0.01; \*\*\* p<0.001. Bonferroni-adjustment applied to coefficient p-values.

Figure 1, below, shows the model calculated from using Structural Equation Modelling with robust standard errors to calculate a cross-lagged panel analysis, controlling for age, income, gender, education and partnership status. Food insecurity (T1) predicted increased depression (T2; r=0.12, p<0.05), anxiety (T2; r=0.14, p<0.05) and food insecurity (T2; r=0.24, p<0.001). Anxiety (T1) predicted higher food insecurity (T2; r=0.1, p<0.05), anxiety (T2; r=0.23, p<0.001), and depression (T2; r=0.18, p<0.001). Depression (T1) predicted increased depression (T2; r=0.1, p<0.05).

Figure 2, below, shows the correlation matrix, respective Spearman rank correlation coefficients and p-values between primary variables averaged at the village level. As shown, Food insecurity (T1) was significantly correlated with anxiety (T1; r=0.72, p=0.01; T2; r=0.67, p=0.02). Anxiety (T2) was significantly correlated with depression (T2; r=0.8, p=0.003).

## DISCUSSION

To support research and response to impacts of climate change on mental health, we analyzed temporal relationships between food insecurity, anxiety, and depression at individual- and village-levels. We found food security predicts subsequent anxiety and depression, controlling for within-variable, within-time, and control variable correlations at the individual-level. At the village-level, we found that aggregated food insecurity (T1) was correlated with significantly higher anxiety (T2). These findings indicate that food insecurity may influence the etiology of anxiety at multiple socio-ecological levels, and depression at the individual-level.

Not only did food insecurity predict higher subsequent depression and anxiety, but also anxiety (T1) predicted higher subsequent food insecurity. As within-time (T2) correlations between food insecurity and anxiety should account for general experiences of worry captured by the food insecurity measure, more research is required to understand psychological mechanisms promoting adaptation to food-insecure environments. As common mental disorders, including anxiety, have previously been shown to undermine productivity in addition to adding direct costs to healthcare expenditures (Christensen et al., 2020). We know of no study that explores how psychological mechanisms, including anxiety, may influence adaptation to food insecure conditions. Understanding and supporting the behavioral, mental, social, and policy conditions that enable positive adaptation to climate change are desperately needed.

That village-level food insecurity was significantly correlated with subsequent anxiety indicates that rural locations where food systems are more adversely affected by climate change may experience subsequently higher levels of anxiety. Mental health promotion efforts should explore the use of existing secondary data on food security, rainfall disruption, and other indicators impacted by climate change to gatekeep whole villages into mental health promotion efforts. The potential for village-level intervention on food security and mental health is indicated by the present study findings. This type of village-level intervention would allow for better understanding of the cultural norms around mental health, anxiety, and depression, as well as food insecurity. Congruent with this village-first strategy, qualitative data indicate people across sub-Saharan Africa view community-resources as a first-line resource for mental health support (Mayston et al., 2020)

As food insecurity, anxiety and depression all significantly improved after 11 weeks in the program, further evaluation of the interventional setting – particularly through a cluster-randomized control trial – is necessary. While the timing between T1 and T2 was relatively short (11 weeks), this period saw food insecurity intensify (Supplemental Figures 1a-b). In contrast, food insecurity, depression, and anxiety all significantly decreased among study respondents in these locations. Whether and how the intervention supports these improvements should be explored in a mechanistic study with robust qualitative data to nuance findings.

Our previous study showed that water insecurity is temporally correlated with anxiety, but food insecurity is not (Goodman et al., 2023). Both studies used different versions of the same scale to measure food insecurity. When we repeated the present study using the shorter scale version, we found results that support our current findings. This suggests that the relationship between food insecurity and anxiety is not related to which version of the scale is used, but to other factors. This ambiguity is in line with previous research on this topic (Pourtmattabed et al., 2020). More research is needed to understand why food insecurity and anxiety are linked in some cases but not others.

### Limitations

The present study has several limitations that should be considered when interpreting the findings. First, the study relied on self-report data, which is susceptible to bias. Individuals may underreport or overreport symptoms of anxiety, depression, and food insecurity due to social desirability or other factors. Additionally, the use of interviewer-administered questionnaires may have introduced interviewer bias, as interviewers may have inadvertently influenced participants’ responses.

Second, the study was conducted in a specific community in Kenya, and the findings may not be generalizable to other populations or settings. Cultural factors and other contextual factors may influence the relationships between food insecurity, anxiety, and depression. Further research is needed to examine these relationships in different populations and settings.

Third, the study was limited by its relatively short follow-up period (11 weeks). Longitudinal studies with longer follow-up periods are needed to better understand the long-term bidirectional effects of food insecurity on mental health.

Fourth, the study did not assess potential mediators or moderators of the relationships between food insecurity, anxiety, and depression. Future research should investigate potential factors that may influence these relationships, such as social support, coping mechanisms, and access to mental health services.

Fifth, community-based microfinance programs tend to recruit substantially higher participation from women than men – as with this study (Brody et al., 2015). As such, future studies should seek to understand barriers for male engagement in these efforts.

Despite these limitations, the present study provides important insights into the complex relationships between food insecurity, anxiety, and depression. The findings underscore the need for holistic interventions that address food security, mental health, and the psychological factors that promote adaptation to food-insecure environments.

## CONCLUSIONS

This study investigated the temporal relationships between food insecurity, anxiety, and depression among adult participants in a community-based empowerment program in Meru County, Kenya. Our findings indicate a complex interplay between these variables at both the individual and village levels. Food insecurity was found to predict subsequent anxiety and depression at the individual level, while village-level food insecurity was correlated with higher anxiety. Additionally, anxiety predicted subsequent food insecurity, suggesting a bidirectional relationship between these variables.

These findings highlight the importance of community-level support and community resilience in promoting mental health in the face of food insecurity. Village-level associations between food insecurity and anxiety suggest that communities with stronger social networks and collective coping mechanisms may be better able to buffer the negative impacts of food insecurity on mental health. Further research is needed to understand the specific factors that contribute to community resilience and to develop interventions that foster these protective factors.

Interventions that address food security, mental health, and the psychological factors that promote adaptation to food-insecure environments are essential for promoting the well-being of individuals and communities in the face of climate change. Community-based approaches that empower individuals and communities to take action to address their own needs are particularly promising.

## Data Availability

All data produced in the present study are available upon reasonable request to the authors

## Supplemental Figures 1a-b

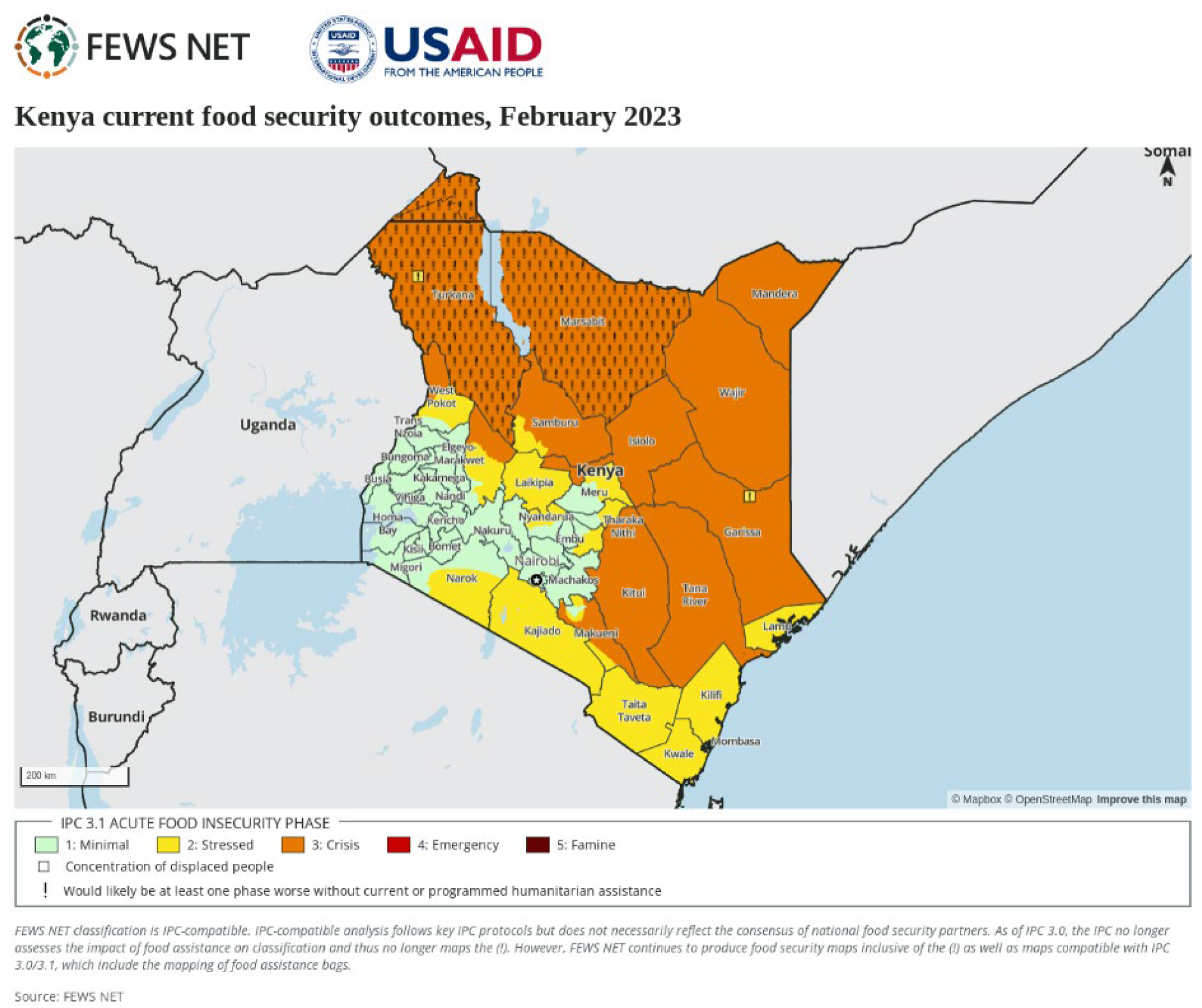

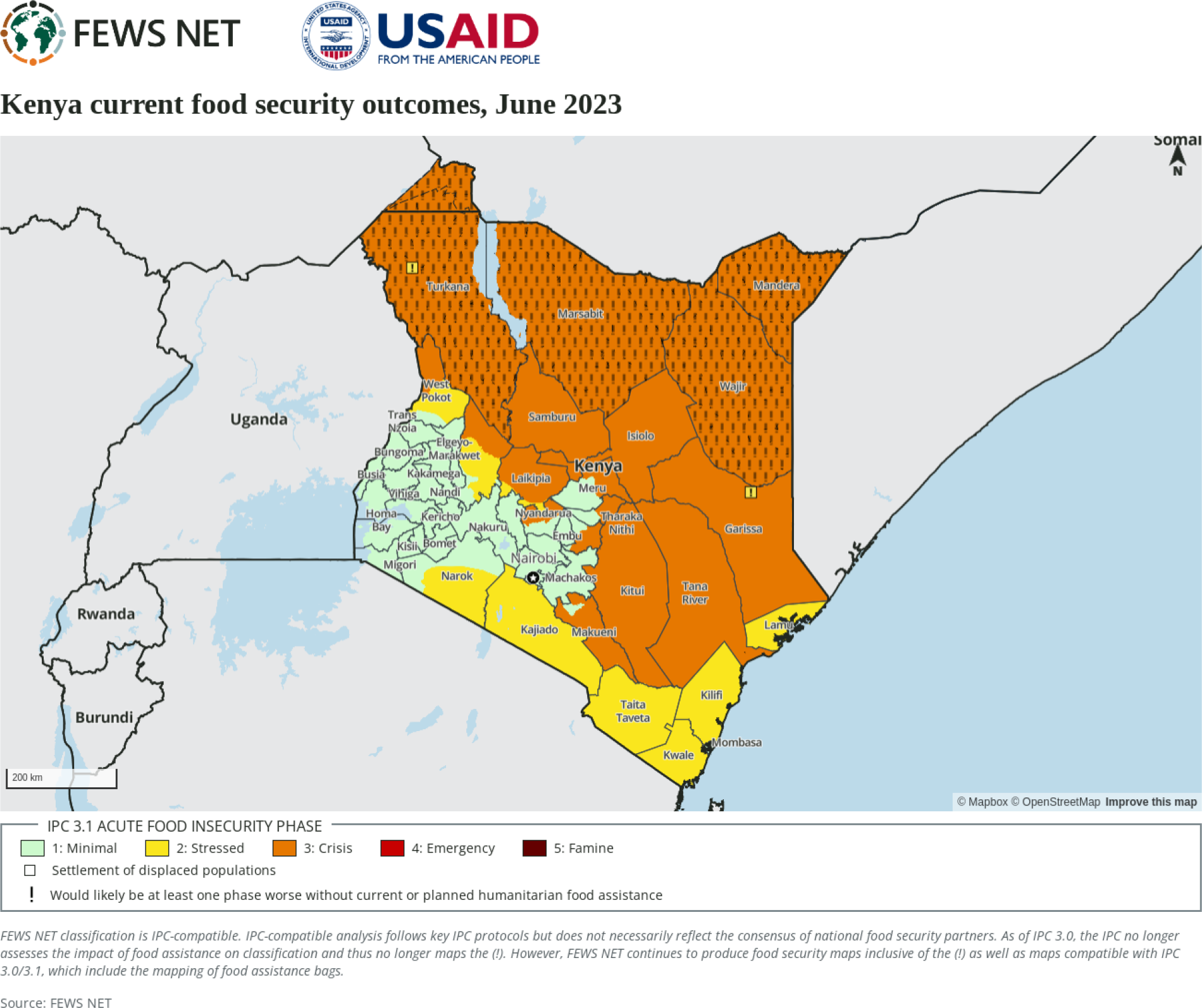

## Study Tables and Figures

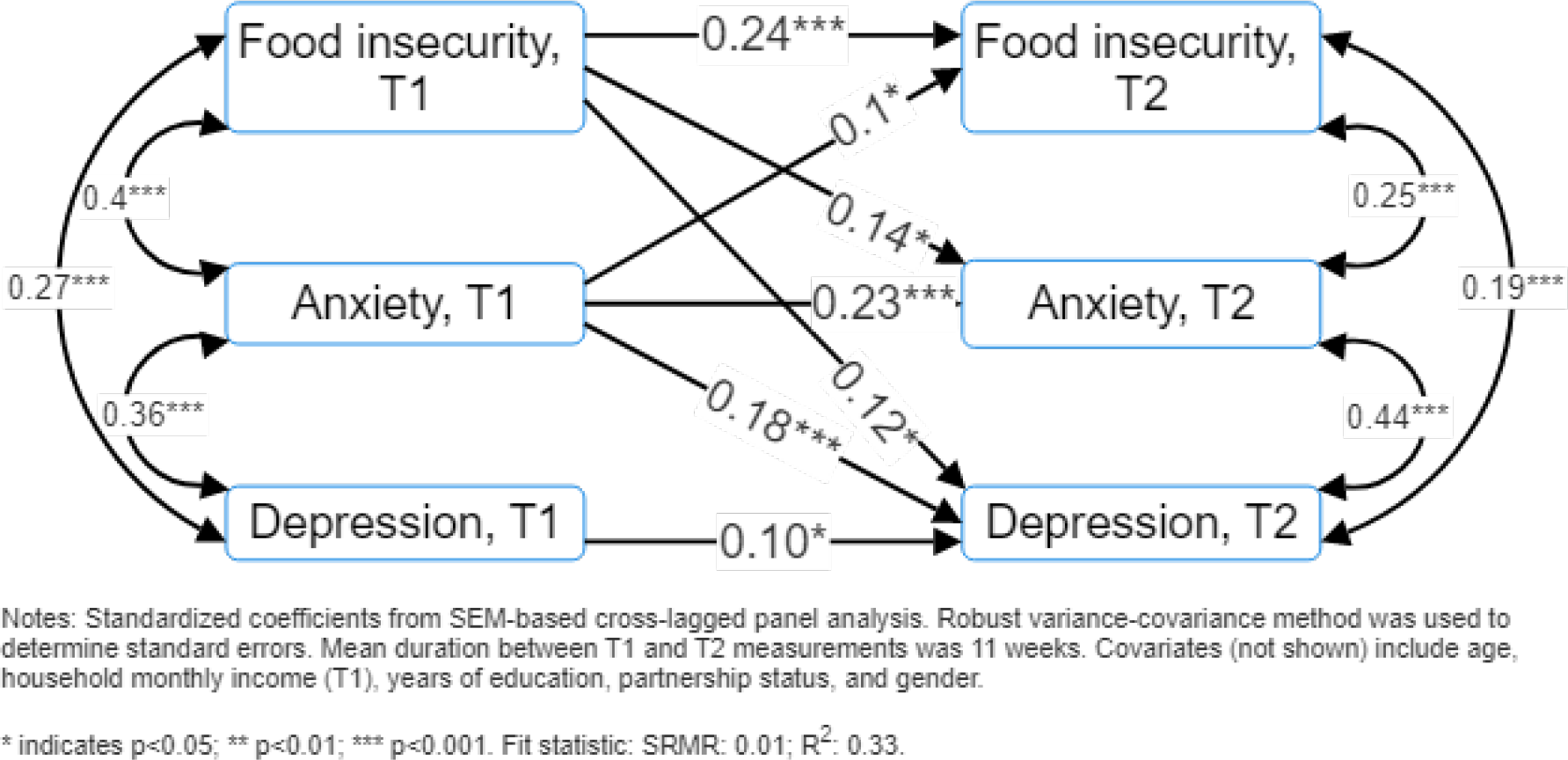

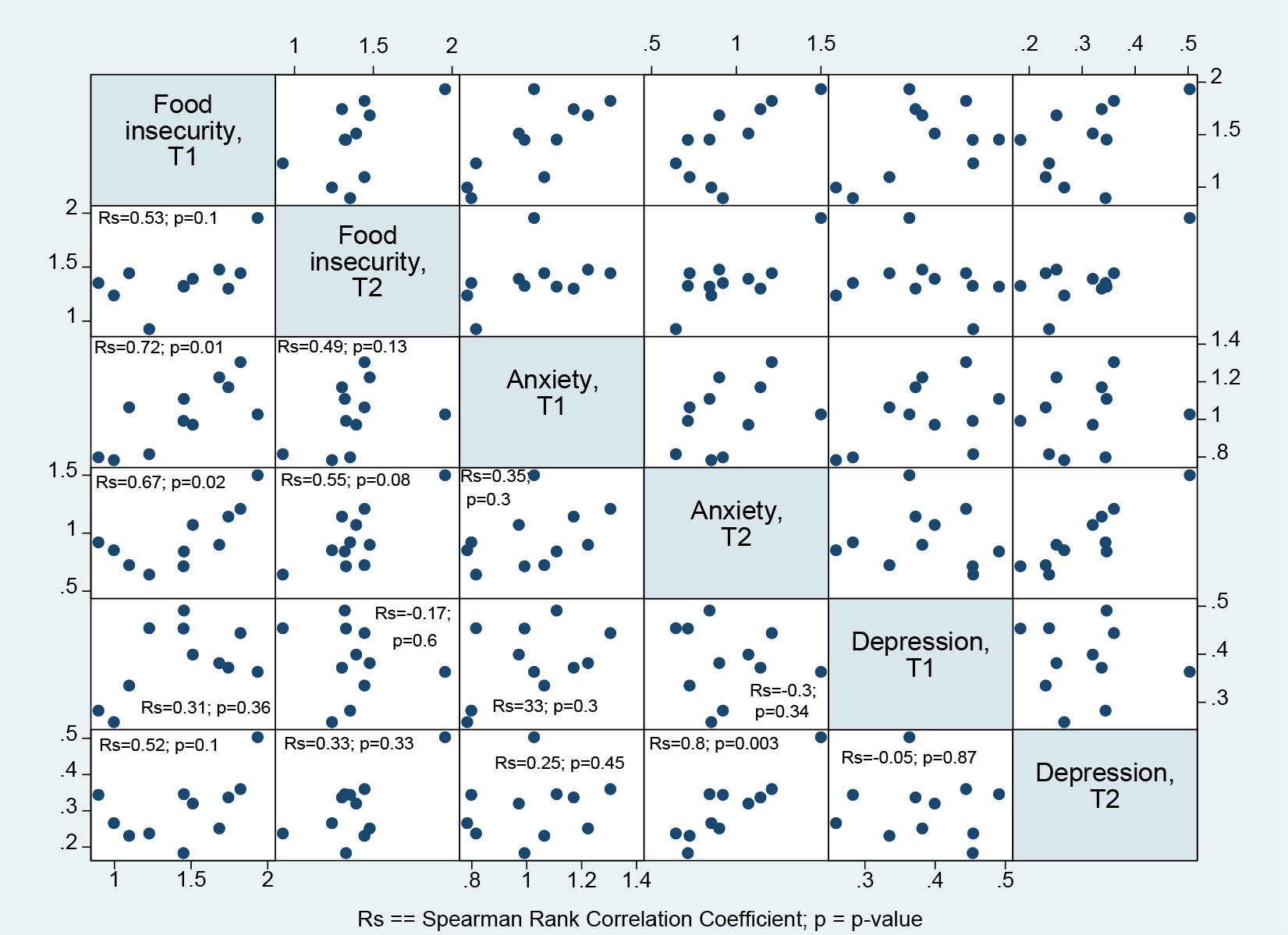

## Notes

### Competing Interest Statement

The authors have declared no competing interest.

### Funding Statement

This study was supported by funding available from the National Institute of Mental Health.

### Author Declarations

The Institutional Review Boards at the University of Texas Medical Branch and the Kenya Methodist University provided approval for this study prior to data collection.

